# Evaluation of nanopore sequencing on polar bodies for routine pre-implantation genetic testing for aneuploidy

**DOI:** 10.1101/2023.04.17.23288670

**Authors:** Anna Oberle, Franziska Hanzer, Felix Kokocinski, Anna Ennemoser, Luca Carli, Enrico Vaccari, Markus Hengstschläger, Michael Feichtinger

**Affiliations:** Wunschbaby Institut Feichtinger, Lainzer Straße 6, 1130 Vienna, Austria; Gene-Test Bioinformatics Solutions, Jakob-Müller-Str. 16, 68623 Lampertheim, Germany

**Keywords:** PGT-A, Polar Body Biopsy, Nanopore Sequencing, Oxford Nanopore Technology, Cost effective PGT-A

## Abstract

**BACKGROUND:** PGT-A using polar body (PB) biopsy derives a clinical benefit by reducing the number of embryo transfers and miscarriage rates but is currently not cost-efficient. Nanopore sequencing technology opens possibilities by providing cost-efficient, fast sequencing results with uncomplicated sample preparation workflows.

**METHODS:** In this comparative experimental study, 102 pooled PB samples from 20 patients were analyzed for aneuploidy using nanopore sequencing technology and compared with aCGH results generated as part of the clinical routine. Samples were sequenced on a Nanopore MinION machine for up to 9 hours for 6 pooled PB samples. Whole-chromosome copy-numbers were called by a custom bioinformatic analysis software. Automatically called results were compared to aCGH results.

**RESULTS:** Overall, 96/99 samples were consistently detected as euploid or aneuploid in both methods (concordance=97.0%, sensitivity = 0.957, specificity = 1.0, PPV = 1.0, NPV = 0.906). On chromosomal level, concordance reached 98.7%. Chromosomal aneuploidies analyzed in this trial covered all 23 chromosomes with 98 trisomies, and 97 monosomies in 70 aCGH samples.

The whole nanopore workflow is feasible in under 5 hours (for one sample) with maximum time of 16 hours (for 12 samples), enabling fresh PB-euploid embryo transfer. Material cost of 150€/sample possibly enable cost-efficient aneuploidy screening.

**CONCLUSIONS:** This is the first study, systematically comparing nanopore sequencing for aneuploidy of PBs with standard detection methods. High concordance rates confirmed feasibility of nanopore technology for this application. Additionally, the fast and cost-efficient workflow reveals clinical utility of this technology, making PB PGT-A clinically attractive.

## Introduction

With increasing female age, the probability for aneuploid embryos increases and with it the risk of age-related infertility, abortions and children born with aneuploidies (1). Early pregnancy loss and spontaneous abortions are most frequently caused by chromosomal abnormalities of the embryo (2). In contrast to maternal age, paternal age plays a minor role for chromosomal abnormalities (3). Preimplantation-genetic testing for aneuploidy (PGT-A) is used to screen for unbalanced chromosomal distributions before implantation and can therefore reduce miscarriage rates and improve pregnancy and life-birth rates per embryo-transfer, especially in patients with advanced-maternal age (4,5).

DNA analyzed in PGT-A can originate from trophectoderm cells, blastomere cells or polar bodies (PBs). All three sources have advantages and specific limitations. While PB analysis allows the detection of meiotic maternal aneuploidies only, the analysis of trophectoderm biopsy (TEB) and blastomere biopsy can detect embryogenic aneuploidies. However, blastomere biopsy can possibly damage the embryo (6,7) and was shown to provide limited clinical benefit (8) and is therefore not commonly performed anymore. In TEB analysis, which is currently seen as the gold standard for PGT-A, diagnosed trophectoderm aneuploidies might differ from embryogenic material of the inner cell mass due to mosaicisms (9). Embryos classified as aneuploid or mosaic by TEB can lead to euploid, healthy life-births, indicating discordances between inner cell mass and trophectoderm or self-repair mechanisms (10,11).

In contrast, PGT-A using PB biopsy leaves less room for interpretation, since mitotic errors in cell division leading to mosaicisms are not detected here. In a large multicenter randomized clinical trial (‘ESHRE Study into the Evaluation of oocyte Euploidy by Microarray analysis’, ESTEEM trial), PGT-A using PB was shown to significantly reduce the number of embryo transfer needed for live birth, reduce number of cryopreserved embryos and reduce miscarriage rates (12). These findings highlight the clinical benefit for PGT-A using PB. In contrast to PGT-A using TEB, PB biopsy allows fresh embryo transfer and can avoid freeze-all procedures. Additionally, due to legal restrictions for TEB analysis in several European countries, PB biopsy is often the only option and is performed frequently in countries like Germany or Austria (13,14).

However, PB analysis was previously stated to be not cost effective (15,16). Lowering the cost for PB-based PGT-A and PGT-A in general is therefore an important goal to improve patient acceptance and accessibility.

PGT-A is often performed using short-read next-generation sequencing (NGS), which requires high initial investment costs and high running expenses. Third-generation sequencing using nanopores is a novel technology with the potential to perform sequencing in a fast, easy, and cost-effective way, enabling application even in small and less well-financed clinics (17). Continuous development of the technology led to significant improvements in sequencing quality (18,19).

Feasibility of nanopore sequencing for PGT was mainly shown for structural variants and monogenetic disease (20–24), which comes with high costs and is far from clinical routine (22). PGT-A from TEB samples using nanopore sequencing was first demonstrated in a small pilot study (25). A follow-up study compared their optimized nanopore aneuploidy analysis of 52 chorionic villi samples, 50 amniotic fluid samples, 64 placental or fetal tissue samples and 52 TEB samples with standard NGS PGT-A screening (26). Overall, the authors saw very good concordance rates. The optimized nanopore workflow was notably faster (2-6 hours) and more cost-effective (around 50 US$ sequencing cost per sample) compared to traditional NGS screening (26). In a recent study, single and multiple cells as well as 96 TEB samples were analyzed for PGT-A and segmental aneuploidies using nanopore sequencing, again confirming high concordance rates above 95% with commercial kits for PGT-A (27). However, further clinical studies are needed to transfer the technology into clinical PGT-A routine analysis.

In the present study, pooled PBs from 102 oocytes were analyzed using nanopore sequencing and compared to results generated as part of the clinical routine using array comparative genomic hybridization (aCGH). This is the first study that analyses aneuploidy from PB samples using nanopore sequencing technology including detailed cost analysis of the nanopore sequencing workflow.

## Material and Methods

### Study design and sampling

In this comparative study, 102 pooled PB samples from 20 patients treated for infertility between March and December 2022 were prospectively collected and analyzed for aneuploidy using nanopore sequencing technology. Nanopore sequencing results were compared with aCGH results generated as part of the clinical routine. All patients participating in this trial were treated for infertility at ‘Wunschbaby Institut Feichtinger’ (WIF) in Vienna and chose aneuploidy screening of their polar bodies. All patients provided written informed consent. The study was approved by the Ethic Committee of the Medical University Vienna (EK-1249/2022). After oocyte pick-up and fertilization by intracytoplasmic sperm injection (ICSI), both PBs 1 and 2 were taken simultaneously 16 to 18 hours after fertilization and placed in one sterile PCR tube with 2.5 µl of fresh, sterile phosphate-buffered saline (PBS). Opening of the zona pellucida was performed using RI Saturn 5 Active^TM^ (Research Instruments Ltd, UK). PB samples were frozen at −20°C and transported to an external genetic diagnostic laboratory for routine analysis.

### DNA Amplification and reference analysis

PGT-A routine analysis of pooled PBs was performed after the standard GentiSure Pre-Screen Kit for Single Cell Analysis Kit (Agilent Technologies, USA) protocol, including Whole Genome Amplification (WGA) with REPLI-g Single Cell Kit using a phi29 polymerase process (QIAGEN, Germany), Fluorescent Labeling and CGH Microarray. This standard protocol was adapted for PB diagnosis. Around 5µg of WGA aliquots from PB samples were anonymized and used for nanopore sequencing.

### Nanopore Library Preparation

After receiving anonymized WGA DNA samples, quality was confirmed by agarose gel electrophoresis (Thermo Fisher Scientific, USA) and quantity was measured by Qubit 4 fluorometer (Thermo Fisher Scientific, Singapore). Then WGA DNA was purified using gDNA Clean & Concentrator-25 columns (Zymo Research, USA).

As part of the protocol optimization, around 1.5 μg of purified DNA was prepared for some samples by digestion of single-stranded regions using T7 Endonuclease I (New England Biolabs (NEB), UK) to reduce or remove branched structures which were introduced during amplification. DNA was purified using gDNA Clean & Concentrator-25 columns (Zymo Research, USA) and eluted in 50µl nuclease-free water. For library preparation, the Oxford Nanopore Technology (ONT, UK) Kits ‘Ligation Sequencing Kit’ (SQL-LSK109) and ‘Native Barcoding Expansion’ (EXP-NBD104) were used according to manufacturer’s recommendations, as previously tested in a pre-clinical trial (28).

Briefly, 1 - 1.5 µg purified DNA was repaired and end-prepared using NEBNext® FFPE DNA Repair and Ultra™ II End-prep Mix (both from NEB, UK) and purified using DNA Clean & Concentrator-5 columns (Zymo Research). For native barcode ligation, 500 to 700 ng DNA was incubated using ONT Native Barcodes and Blunt/TA Ligase Master Mix (NEB) and purified using DNA Clean & Concentrator-5 columns (Zymo Research). Purified, barcoded DNA samples for one sequencing run (between 4 to 8 samples, see Supplementary Table S1) were pooled equimolar in one tube and ligated to AMII adapters (ONT) with NEBNext® Quick Ligation Reaction Buffer and Quick T4 DNA Ligase (both NEB). For the final clean-up, 0.5x SPRIselect beads (Beckman Coulter, USA) were utilized to ensure elimination of small DNA fragments and washed with Short Fragment Buffer (ONT) before elution.

### Nanopore sequencing

The sequencing-ready library was primed and loaded on a MinION R9.4 flow cell (FLO-MIN106D, ONT) according to manufactures recommendation using 25-50 fmol DNA library. Sequencing was performed on MinION Mk1C with simultaneous base calling using standard base calling settings and quality threshold (minimum Q-score) of 8. Sequencing time was adapted according to number of samples pooled and quality of the flow cell (pores available) with average sequencing time of around 9 hours for 6 pooled PB samples resulting in a median number of 105,670 reads (after pre-processing, stdev= 76,444) and 210 Mbases (stdev=100) generated per sample (detail sequencing constitution, time and output in Supplementary Table S1).

### Data analysis

Nanopore’s *MinKNOW* software system controls the raw data acquisition, performing basecalling and demultiplexing with *Guppy* as well as FASTQ file creation. Following on, a data analysis pipeline was applied, developed for long read-based PGT-A data using standard bioinformatics tools and custom software scripts. The specific processing steps are described in the following.

Primary data analysis consisted of read cleaning with *Porechop* (Porechop, RRID:SCR_016967) and *Nanofilt* (29), alignment to the human genome GRCh38 with *minimap2* (30) and creation of BAM files with *samtools* (31). Secondary processing steps made use of the *QDNAseq* R software package (32) and include binning of reads using a custom 500 kb bin matrix, GC content and mappability correction, median and reference normalisation, smoothing, segmentation and copy-number calling. A set of five samples were used as a combined reference to capture experiment-specific bias. They represented the first few samples of the study with a representative profile and without whole-chromosome copy-number changes as assessed by aCGH. Further processing was performed to filter and summarize the data, producing a report with genome-wide copy-number plots, noise measurements and automatically detected changes per chromosome and sample. Samples with more than 3 aneuploid chromosomes were classified as ‘multiple’.

As quality measures of the nanopore sequencing samples, MAPD (Median of the Absolute values of all Pairwise Differences) and noise (median standard deviation of normalised read counts within each segment) values are reported for all chromosomes as well as for each sample and automatically plotted in the chromosomal distribution plots. Samples resulting in a total noise value greater than 0.6 and MAPD value greater than 1.7 were excluded from the comparison (Supplementary Table S1).

## Results

In this study, 102 pooled PB samples from 20 patients were analyzed for PGT-A by nanopore sequencing and compared to clinical routine PGT-A analysis by aCGH. All samples were successfully amplified while a negative control (culture medium) for each batch showed no amplification and aCGH was performed for all 102 PB samples. WGA aliquots were anonymized and used for nanopore sequencing. Schematic study design is shown in Figure 1.

**Figure 1:**
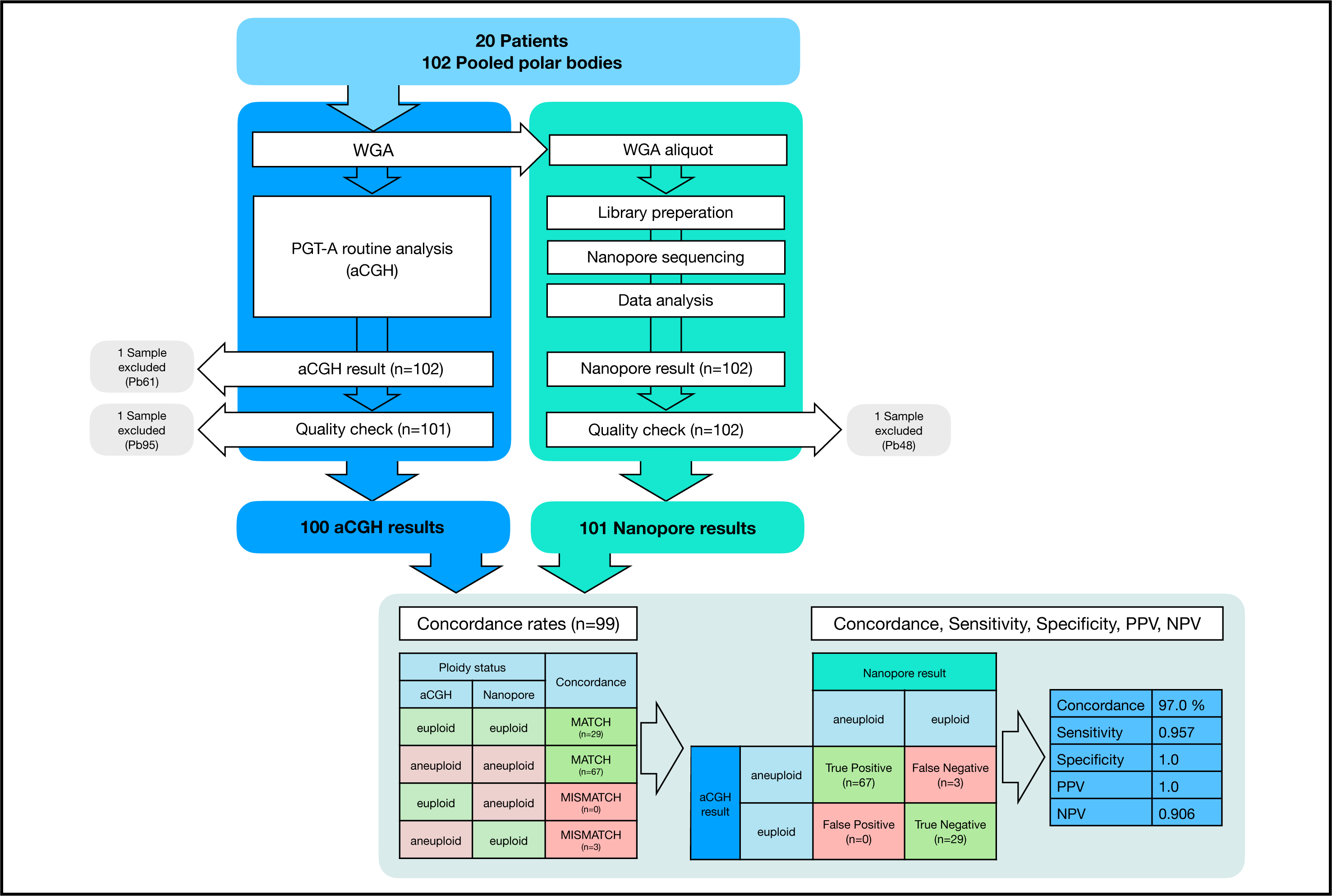
Study design including concordance rates found in this study. WGA: whole-genome amplification; PGT-A: preimplantation-genetic testing for aneuploidy; aCGH: array comparative genomic hybridization; PPV and NPV: positive and negative predictive value.

**Figure 2:**
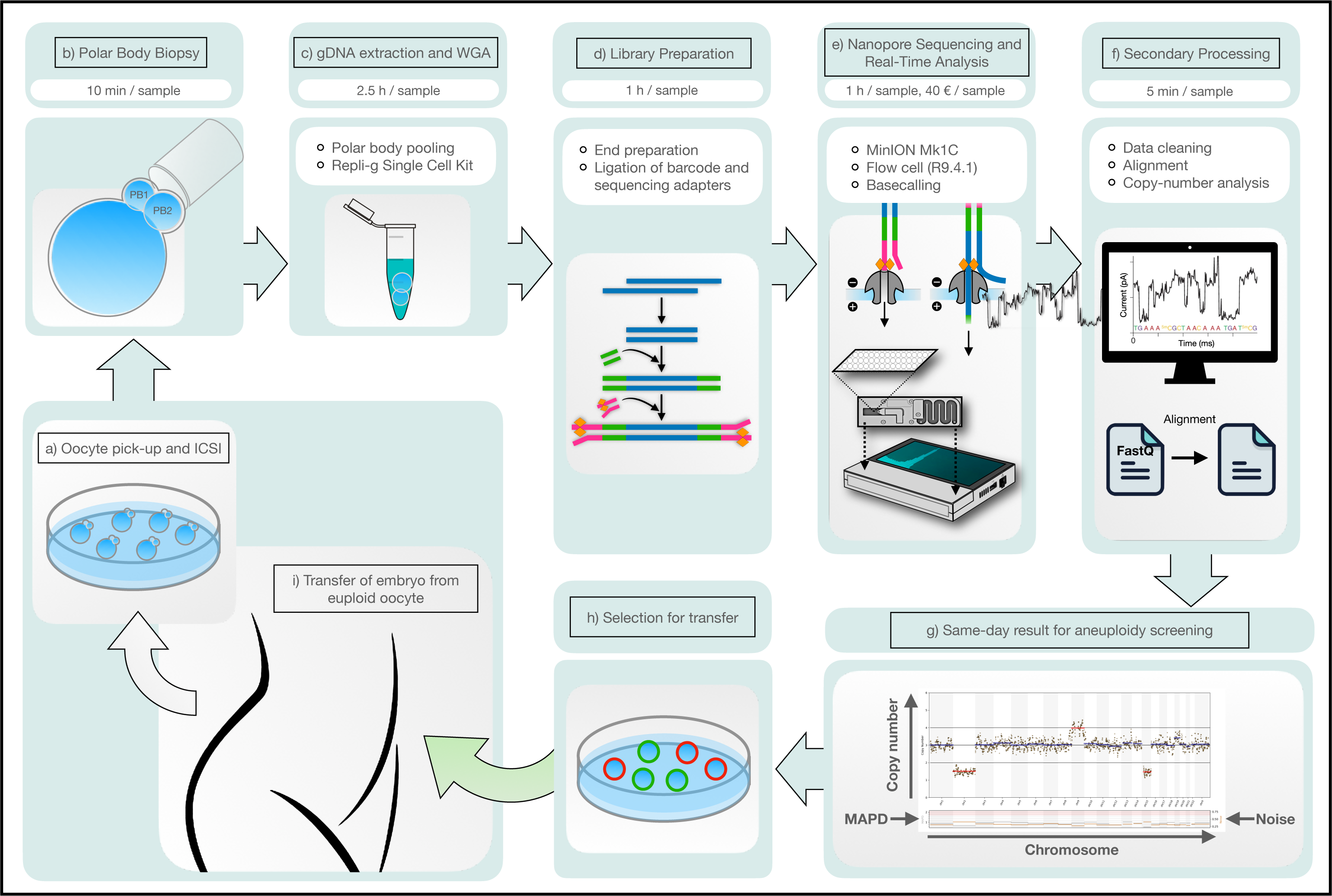
Schematic overview of the nanopore sequencing workflow, including time measures for individual procedures, embedded in the clinical PGT-A workflow. Prorate sequencing time per sample was calculated according to medium sequencing time for 6 pooled samples. Separate sequencing of one sample can be shorter. ICSI: intra-cytoplasmatic sperm injection; PB: polar body; WGA: whole-genome amplification.

### Quality control and genome coverage for nanopore sequencing workflow

In total three samples were excluded from the comparison. Two samples (PB61, PB95) were excluded due to failed or uninterpretable aCGH (2 %) and one sample (PB48, 1%) was excluded due to failed quality check after nanopore sequencing with noise and MAPD values below quality threshold.

Data analysis from nanopore sequencing results revealed medium sequencing output of 210 Mbases and a median of 105,670 reads per sample (after pre-processing, 70 to 650 Mbases and 32,924 to 374,841 reads). A median of 3% (88 Mbases) of the genome was covered by one or more reads (between 0.9 – 9.3%). With the 0.5 Mbases bin matrix applied in the analysis, a smoothing window of 6 bins and a calling limit of 20 bins, the application has a theoretical resolution of around 10 Mbases. This is comparable with the common resolution for PGT-A using aCGH or NGS (6-10 Mbases) (33). Average Q-score of all nanopore-sequenced samples was 8. Quality was additionally verified in the bioinformatic data analysis pipeline by noise and MAPD values. Average noise of all samples was 0.428 (between 0.230 and 0.724) and average MAPD was 1.164 (between 0.720 and 1.809). All noise and MAPD values are listed in Supplementary Table S1 and realtionship between both quality measures is visualized in Supplementary Figure S1.

### Comparison of PGT-A results from nanopore sequencing with routine aCGH results

In total, 99 pooled PB samples for PGT-A screening were utilized in this study for comparison of aCGH with nanopore sequencing workflow. From these, 29 were detected euploid (29.3 %) and 70 were detected aneuploid (70.7 %) by aCGH reference. This is expected, given the patient cohort with medium female age of 40.5 years. Nanopore sequencing analysis detected 32 samples as euploid and 67 as aneuploid resulting in 97.0 % sample-level concordance and a sensitivity of 0.957, specificity of 1.0 with a positive predictive value (PPV) of 1.0 and a negative predictive value (NPV) of 0.906 (Figure 1). The results of all aCGH and nanopore sequencing analyses are listed in Table 1. Three samples detected as aneuploid using aCGH revealed an euploid chromosomal pattern using nanopore sequencing. All three samples showed a single aneuploid chromosome (+22, +19, −16, respectively) by aCGH and these elevations or reductions were visible but below threshold at nanopore sequencing, as listed in Supplementary Table S1.

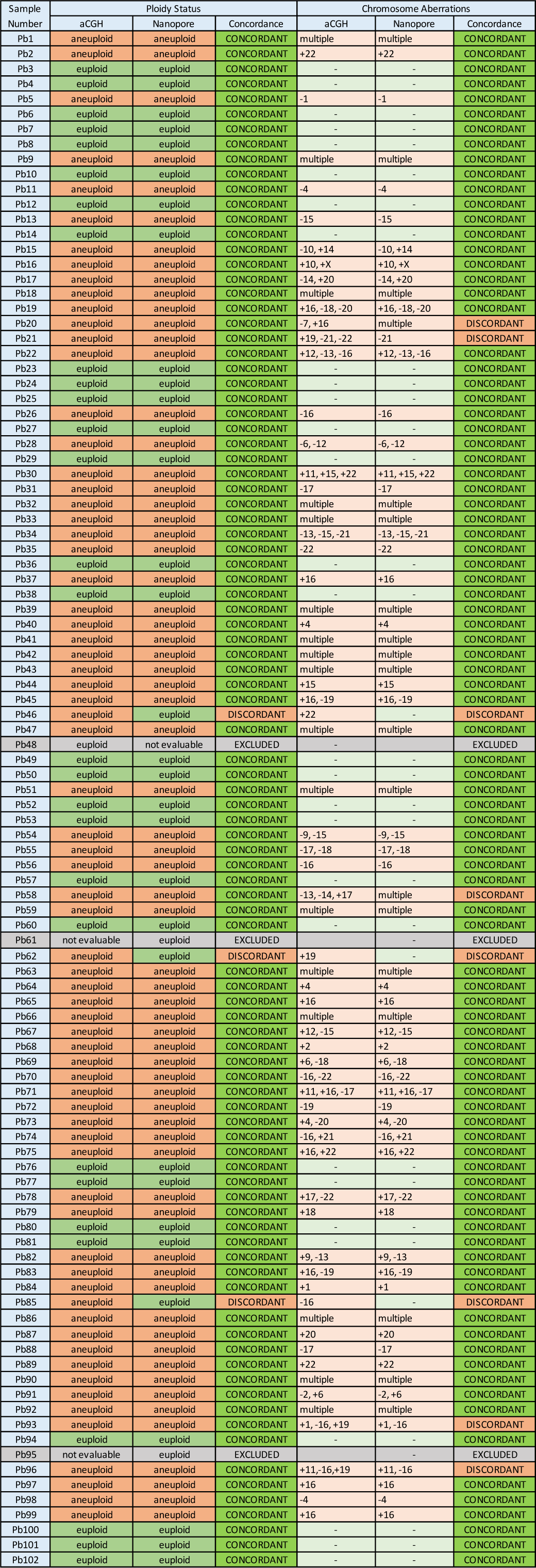

In total, aCGH analysis detected 195 aneuploid chromosomes in the 99 samples, with 98 trisomies and 97 monosomies covering all 23 chromosomes, shown in Figure 3. Counting the exact match (all chromosomes concordant but assigning “multiple” to karyotypes with more than three aberrations), we found 91.9% (karyotype-level) concordance (91 of 99 samples showing exact match), as listed in Table 1. On chromosomal level, we found 98,7% concordance (2244 of 2273 chromosomes were concordantly identified as normal or abnormal). Sensitivity was 0.901, specificity 0.995, PPV 0.945 and NPV 0.991 per chromosome. Segmental changes were not addressed in this study. Example chromosomal distribution plots of euploid and aneuploid detected samples, which are automatically created from bioinformatic data analysis pipeline after nanopore sequencing are shown in Figure 4.

**Figure 3:**
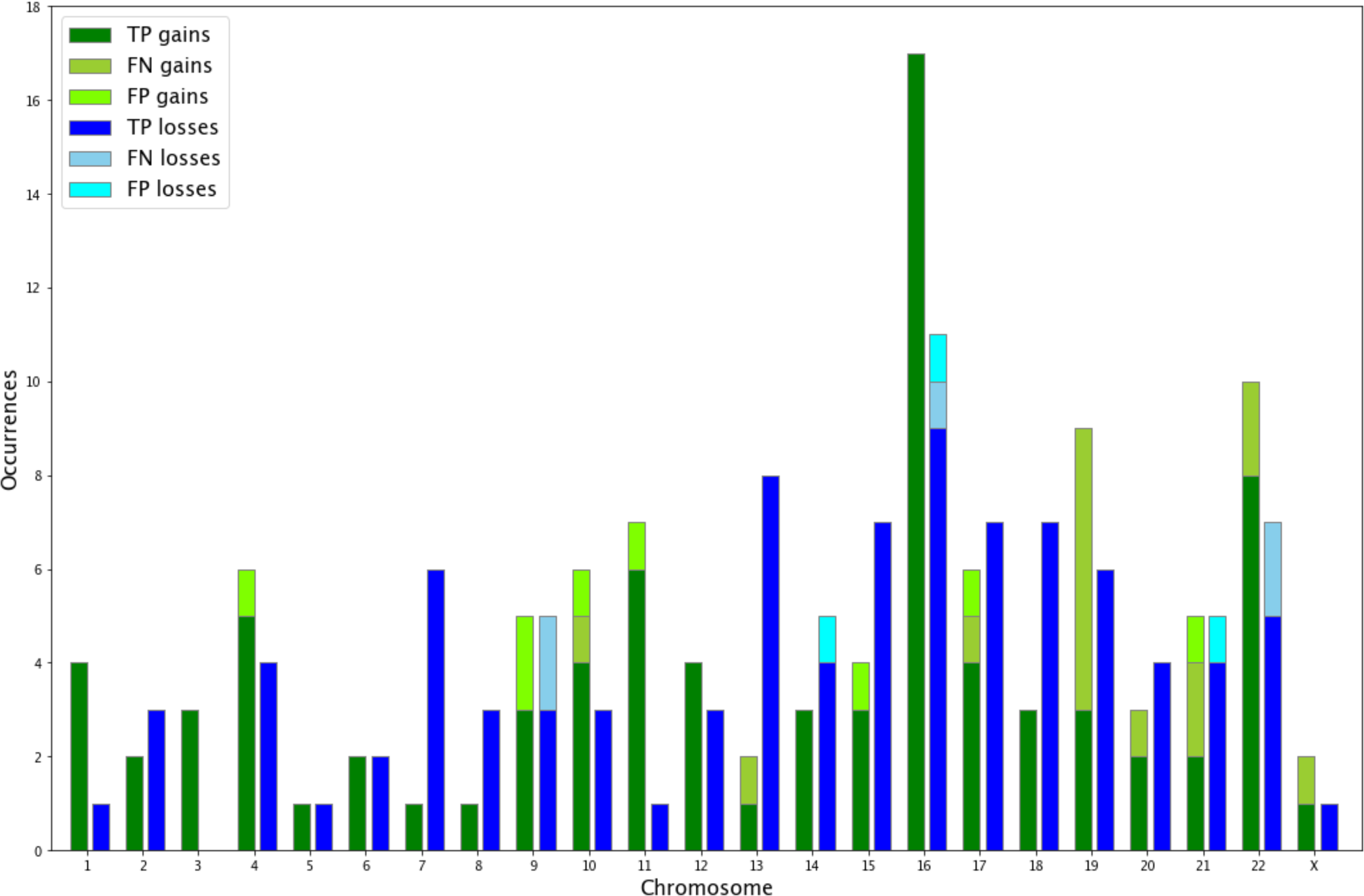
Occurrence of whole-chromosome aneuploidies in all 99 pooled PB samples. TP: true positive, FN: false negative, FP: false positive.

**Figure 4:**
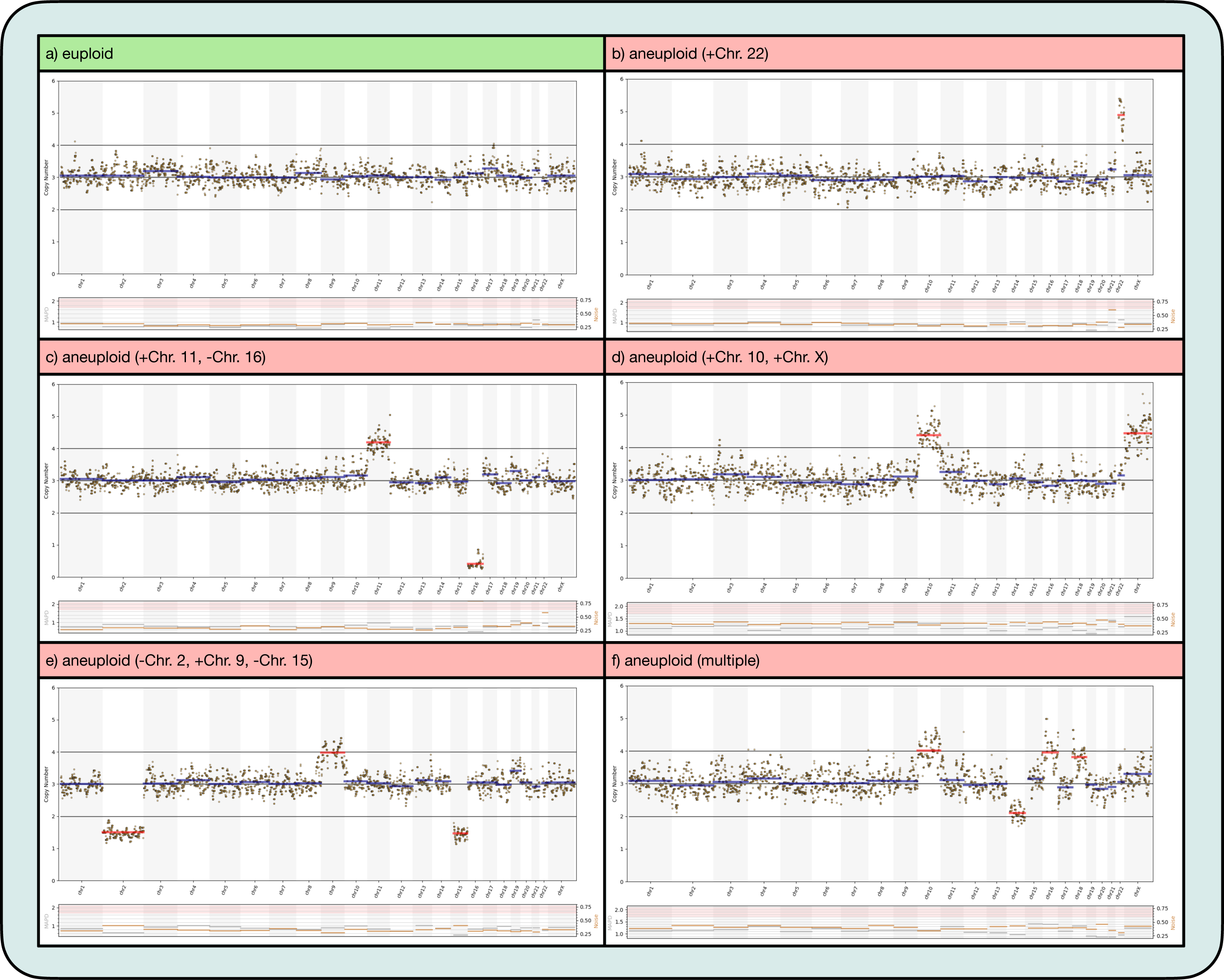
Example chromosomal distribution plots from automatically generated nanopore sequencing workflow. Quality values noise and MAPD are mapped for each chromosome and thresholds for the total sample (0.6 and 1.7, respectively) are highlighted in red. MAPD: Median of the Absolute values of all Pairwise Differences; noise: median standard deviation of normalised read counts within each segment.

To assist the physicians and patients in the process of decision-making, a further critical evaluation of euploid-detected PBs can be performed and marginal/borderline elevated or reduced chromosomes can be mentioned together with the euploid result in the clinical report. Here, we performed manual revision of all euploid-detected samples after nanopore sequencing and the additional information about slightly elevated or reduced chromosomes can be found in Supplementary Table S1.

### Endonuclease digestion step

As part of this study, we evaluated the importance of an endonuclease digestion step. 28 samples were run in parallel with and without endonuclease digestion step. While samples with a high signal to noise ratio show matching results, the additional digestion clearly reduces noise and improves calling for challenging cases. Reduction of average noise and MAPD values, as well as improvements of sequencing-output per time is shown in Supplementary Figure S2. Detailed comparison of sequencing results with and without endonuclease digest is listed in Supplementary Table S2. For the calculation of concordance between aCGH and nanopore sequencing, results after Endonuclease digest were utilized for all these samples. For all new investigations we recommend adding this step.

### Detailed time and cost calculation

To evaluate if nanopore sequencing for PGT-A can be utilized in clinical routine, we performed a detailed time and cost analysis for our nanopore sequencing workflow, which is listed in Supplementary Tables S3 and S4. The nanopore sequencing workflow used in this study starting from amplified DNA until ploidy result is feasible in one and a half hours with one sample being sequenced separately. For 12 samples being sequenced in parallel, the workflow is feasible in around 12 hours (Supplementary Table S3). Amplification of the pooled PB additionally take between 1.5 hours and 4.5 hours per sample, depending on the WGA kit. In our study, WGA was performed using REPLIg Single Cell Kit (QIAGEN) with a 2.5 h amplification protocol.

Material cost for the whole workflow, including WGA, sample preparation, sequencing and data analysis range from 100€ to 220€ per sample. A detailed list of time and cost calculation for different scenarios is shown in Supplementary Table S3 and S4. Initial investment cost for nanopore sequencing can be neglected because prices for nanopore starter packages, where MinION sequencing machines are included cover material costs.

## Discussion

The present study is the first study that systematically compares PB PGT-A analysis using nanopore sequencing with routine aCGH analysis. The concordance rate between both technologies found in this study was 97% for euploid/aneuploid sample decision and 99% on chromosomal level. These concordance rates are high, given the different nature of the methods and the delicate sample type of PB biopsy. Similar studies comparing different technologies for PGT-A analysis received slightly higher concordance rates (26,34–36). However, genetic material utilized in these studies was originated from TEB, which comprises significantly more genetic starting material than PB biopsy, resulting in more stable and uniform whole genome amplification. From 102 PB samples analyzed with nanopore sequencing in our study, 101 samples passed the quality threshold, indicating a high reliability of the technology.

Nanopore sequencing has several advantages, compared to aCGH or NGS analysis, including but not limited to the fast turnaround time, inexpensive sequencing, as well as small initial investment costs needed. These advantages might lead to cost-efficient diagnostics for clinical applications. Time and cost of nanopore sequencing for clinical reproductive healthcare has been reported in several publications, but detailed statements on how these number can be achieved are often not stated (20,25,26,37). Here, we provide detailed insight in material costs and preparation, sequencing, and data analysis time for different scenarios (Supplementary Table S3 and S4).

Another advantage of nanopore sequencing compared to aCGH or NGS is the high flexibility of the sequencing method. Sequencing data as well as quality measures of the data generated, and the flow cell health can be monitored in real-time during nanopore sequencing. If more data is required or quality of the library is poor, sequencing can easily be prolonged or repeated on the same flow cell until enough data is generated. This can reduce the necessity to repeat uninterpretable results and can reduce prolonged time-to diagnosis, increasing the possibility for same-cycle transfer of euploid embryos, leading again to a cost reduction by reducing freeze-all procedures and hormonal treatment for the embryo transfer, possibly leading to a shorter time-to-pregnancy.

A possible limitation of the study derives from the reference method. In a comparable study using nanopore sequencing in comparison with standard clinical detection methods (NGS, aCGH or fluorescence in situ hybridization, FISH), discordant aneuploidy results were re-examined and the initial nanopore sequencing results were confirmed in these samples, showing that even established clinical testing methods like FISH or aCGH might result in misdiagnosis (26). Misdiagnosis of routine aCGH PGT-A analysis has also been shown in a study comparing aCGH with NGS results (35). In the present study using material from pooled PBs, re-examination of aCGH was not possible due to limited sample resource.

For some samples in this study, the reference result remained unclear, due to increased noise and limited resolution of the aCGH method. In particular, the small chromosomes show higher variability in both platforms. Discordant results could originate from different thresholds defined in aCGH and nanopore sequencing, different analysis bias and bias reduction (in the nanopore workflow, bias was reduced through binning, GC content and mappability correction, median and reference normalisation), or different genome covargae and resolution. Although the reference method has been extensively validated (5) and is applied in clinical routine since 2015, the utilization of pooled PBs, analyzed with aCGH might come with some technical limitations, including limited resolution and the necessity for manual revision. While the assessment and quantification of the nanopore sequencing results is clearer, manual review is still recommended and sometimes required. A larger study with follow-up data or a clinical non-selection trial would be beneficial for increased confidence.

Utilizing pooled PBs, compared to analyzing PB 1 and 2 separately, requires more sensitive detection methods, since variations from normal euploid chromosomes depict less distinct in pooled PBs. However, if properly validated (5), pooling PBs can considerably reduce the costs and the workload of the analysis.

Independent of the technology, the analysis of chromosomal material is complicated by different characteristics of human chromosomes (38). The short chromosomes 19, 20, 21, 22 and Y provide limited material which leads to an increased variability in measurements. The GC-richest chromosomes 16, 17, 19 and 22 often display increased levels of noise. Also, chromosomes with heterochromatic polymorphisms, like chromosomes 1, 9 and 16 differ due to normal, benign variations in heterochromatin-content, which can be visualized by microscopy but are hard to interpret in sequencing-based methods or aCGH (39). In our study, the chromosomes 9, 19, 21 and 22 showed most discordances (Figure 3).

Although manual review of the chromosomal distribution of each sample is recommended, our data analysis pipeline automatically generates the sequencing plot with significantly abnormal chromosomes being highlighted and quality measures (noise and MAPD) are plotted for each chromosome. The karyotype is reported automatically according to the optimized bioinformatic pipeline. This automation together with the clear and easy-to-read karyotype plot makes the nanopore sequencing data very easy to understand and interpret. Interpretation of the results does not require years of expert knowledge and experience, which is often required for the interpretation of aCGH plots. The automated analysis pipeline is not dependent on subjective decision-making of the responsible geneticist or physician. Nevertheless, following the clinical practice, we additionally performed manual review of all euploid-detected samples (Supplementary Table S1). This additional annotation about slight chromosomal aberrations does not change the automatically reported result (euploid) but can be provided as additional information in the medical report. This report can facilitate the embryo prioritization and decision-making for the physician and the patients as part of their fertility treatment.

Taken together, pooled PB nanopore sequencing revealed high concordance rates compared to conventional aCGH with minimized time and financial resources required.

## Data Availability

The data files (raw and adjusted read counts per genomic region) can be freely accessed by other researchers for further analysis at https://www.gene-test.com/oberle-et-al-2023

## Acknowledgements

We would like to thank all patients participating in this study. We also thank all staff members from HLN Genetic GmbH for providing the reference samples.

## Notes

### Competing Interest Statement

The authors have declared no competing interest.

### Funding Statement

This study was funded by the Dr Wilfried Feichtinger GmbH

### Author Declarations

The study was approved by the ethical committee of the medical university of Vienna

### Summary of Updates

Latest important publications to the topic have been implemented in the manuscript and it has been formatted for publication.

