## Supplementary material for "Evaluation of nanopore sequencing on polar bodies for routine pre-implantation genetic testing for aneuploidy": Suppl. Figure and Table Legends

**Supplementary Figure and Table Legends:**

**Supplementary Figure S1: Relation of noise and MAPD quality values from all 102 PB samples analyzed with nanopore sequencing**. MAPD: Median of the Absolute values of all Pairwise Differences;  noise: median standard deviation of normalised read counts within each segment.

**Supplementary Figure S2: Comparison of nanopore sequencing workflow with and without Endonuclease I digest**. EN: Endonuclease I digest.

**Supplementary Table S1: Detailed list of Nanopore sequencing results in comparison with aCGH results.** All samples, that were digested with Endonuclease I after WGA are marked as EN. MAPD: Median of the Absolute values of all Pairwise Differences;  noise: median standard deviation of normalised read counts within each segment; TP: true positive; TN: true negative; FP: false positive; PN: false negative

**Supplementary Table S2: Outcome measures of nanopore sequencing samples with and without Endonuclease I digestion step**.

**Supplementary Table S3: Calculation of time and cost of Nanopore sequencing workflow for polar body PGT-A**.

**Supplementary Table S4: Detailed list of costs for consumable material for nanopore sequencing workflow**. All prices were obtained in December 2022 from the respective supplier websites with Austrian-based settings.
