## Supplementary figures and images for "Evaluation of nanopore sequencing on polar bodies for routine pre-implantation genetic testing for aneuploidy"

### Supplementary Figure S1

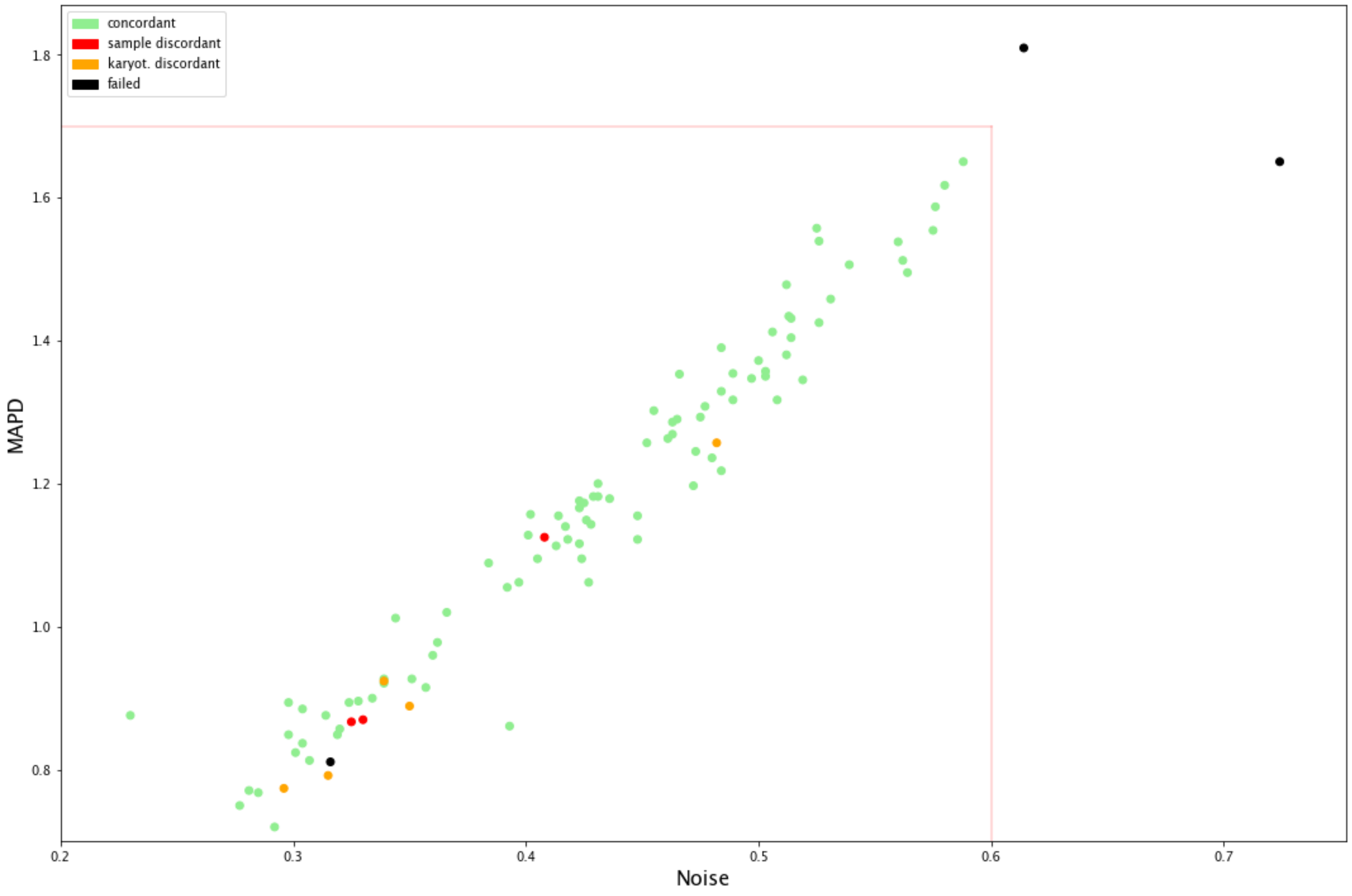

### Supplementary Figure S2

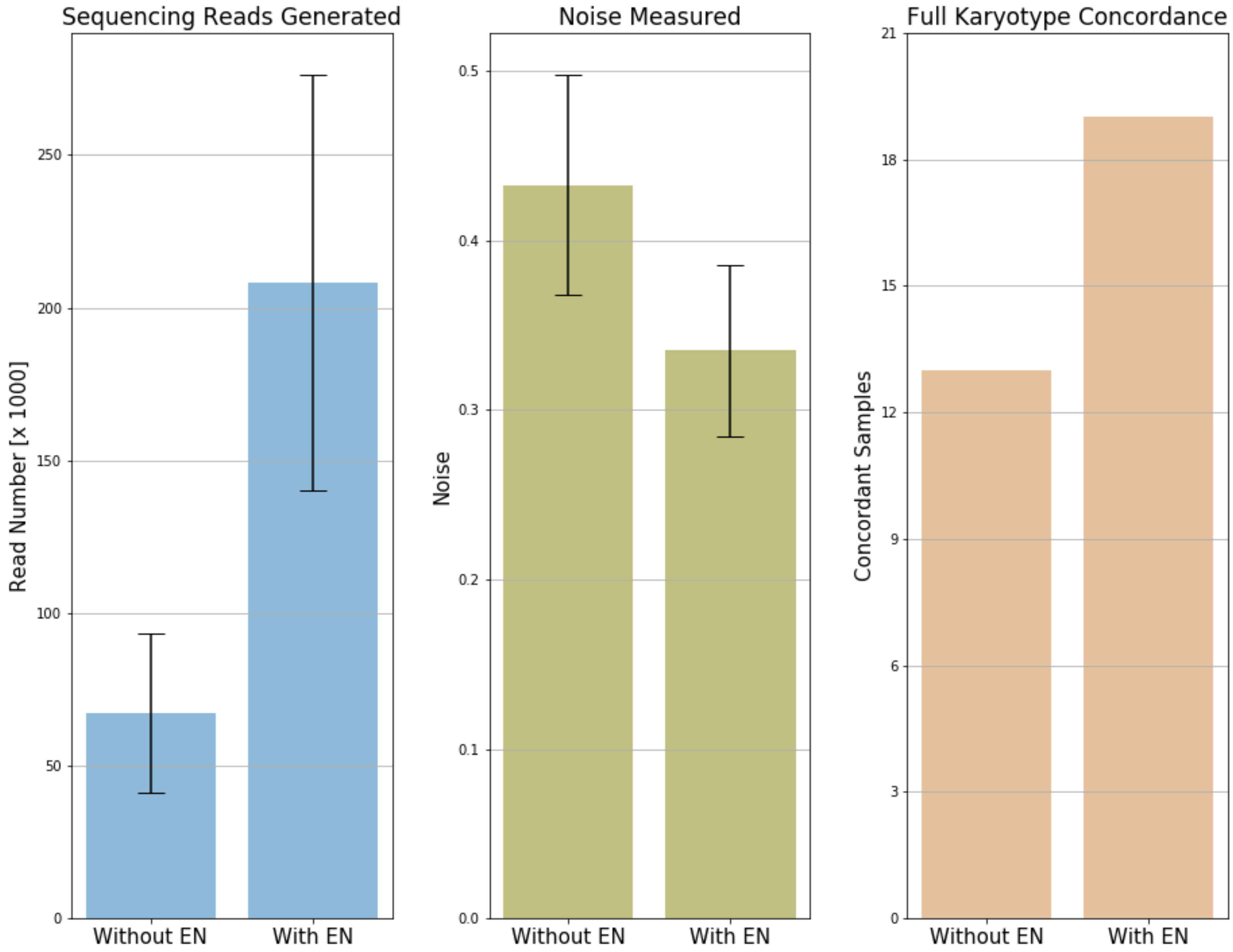
