## Supplementary Table S1 for "Evaluation of nanopore sequencing on polar bodies for routine pre-implantation genetic testing for aneuploidy"

| Sample Number | Conc. after WGA (ng/µl) | Library Preperation | | Nanopore Sequencing | | Output | | | | Ploidy Status | | | Sample-Level Assessment (euploid/aneuploid) | | | | Chromosome Aberrations | | | manual evaluation of euploid nanopore results | Chromosome-Level Assessment (individual chromosomes affected) | | | |
|---|---|---|---|---|---|---|---|---|---|---|---|---|---|---|---|---|---|---|---|---|---|---|---|---|
| | | Endonclease digest | Barcode | Run | Time / Sample (min) | Yield [Mbases] | Raw Reads | noise | MAPD | aCGH | Nanopore | Concordance | TP | TN | FP | FN | aCGH | Nanopore | Concordance | | TP | TN | FP | FN |
| Pb1 | 400 | no | 1 | 1 | 90 | 130 | 121.912 | 0,360 | 0,960 | aneuploid | aneuploid | CONCORDANT | 1 | 0 | 0 | 0 | multiple (+2, -9, -15, -16) | multiple (+2, -9, -15, -16) | CONCORDANT | | 4 | 19 | 0 | 0 |
| Pb2 | 239 | no | 2 | 1 | 90 | 133 | 138.932 | 0,339 | 0,921 | aneuploid | aneuploid | CONCORDANT | 1 | 0 | 0 | 0 | +22 | +22 | CONCORDANT | | 1 | 22 | 0 | 0 |
| Pb3 | 283 | no | 3 | 1 | 90 | 104 | 94.894 | 0,304 | 0,885 | euploid | euploid | CONCORDANT | 0 | 1 | 0 | 0 | - | - | CONCORDANT | ok | 0 | 23 | 0 | 0 |
| Pb4 | 264 | no | 4 | 1 | 90 | 111 | 104.648 | 0,314 | 0,876 | euploid | euploid | CONCORDANT | 0 | 1 | 0 | 0 | - | - | CONCORDANT | ok | 0 | 23 | 0 | 0 |
| Pb5 | 447 | no | 5 | 2 | 70 | 115 | 104.473 | 0,392 | 1,055 | aneuploid | aneuploid | CONCORDANT | 1 | 0 | 0 | 0 | -1 | -1 | CONCORDANT | | 1 | 22 | 0 | 0 |
| Pb6 | 347 | yes | 1 | EN4 | 90 | 170 | 117.144 | 0,339 | 0,927 | euploid | euploid | CONCORDANT | 0 | 1 | 0 | 0 | - | - | CONCORDANT | ok, (+22?) | 0 | 23 | 0 | 0 |
| Pb7 | 323 | no | 7 | 2 | 70 | 108 | 90.215 | 0,298 | 0,849 | euploid | euploid | CONCORDANT | 0 | 1 | 0 | 0 | - | - | CONCORDANT | ok | 0 | 23 | 0 | 0 |
| Pb8 | 323 | no | 8 | 2 | 70 | 154 | 133.237 | 0,351 | 0,927 | euploid | euploid | CONCORDANT | 0 | 1 | 0 | 0 | - | - | CONCORDANT | ok, (+20?) | 0 | 23 | 0 | 0 |
| Pb9 | 309 | no | 9 | 2 | 70 | 62 | 50.466 | 0,452 | 1,257 | aneuploid | aneuploid | CONCORDANT | 1 | 0 | 0 | 0 | multiple (+7, +10, +20, +21, -22) | multiple (+7, +10, +21, -22) | CONCORDANT | | 4 | 18 | 0 | 1 |
| Pb10 | 303 | yes | 1 | EN2 | 100 | 178 | 189.572 | 0,324 | 0,894 | euploid | euploid | CONCORDANT | 0 | 1 | 0 | 0 | - | - | CONCORDANT | ok | 0 | 23 | 0 | 0 |
| Pb11 | 307 | no | 11 | 3 | 90 | 84 | 67.208 | 0,423 | 1,176 | aneuploid | aneuploid | CONCORDANT | 1 | 0 | 0 | 0 | -4 | -4 | CONCORDANT | | 1 | 22 | 0 | 0 |
| Pb12 | 307 | yes | 2 | EN2 | 100 | 245 | 274.533 | 0,281 | 0,771 | euploid | euploid | CONCORDANT | 0 | 1 | 0 | 0 | - | - | CONCORDANT | ok | 0 | 23 | 0 | 0 |
| Pb13 | 302 | no | 3 | 3 | 90 | 64 | 48.057 | 0,466 | 1,353 | aneuploid | aneuploid | CONCORDANT | 1 | 0 | 0 | 0 | -15 | -15 | CONCORDANT | | 1 | 22 | 0 | 0 |
| Pb14 | 299 | yes | 7 | EN3 | 100 | 144 | 190.505 | 0,328 | 0,896 | euploid | euploid | CONCORDANT | 0 | 1 | 0 | 0 | - | - | CONCORDANT | ok | 0 | 23 | 0 | 0 |
| Pb15 | 440 | no | 5 | 4 | 90 | 100 | 72.449 | 0,423 | 1,166 | aneuploid | aneuploid | CONCORDANT | 1 | 0 | 0 | 0 | -10, +14 | -10, +14 | CONCORDANT | | 2 | 21 | 0 | 0 |
| Pb16 | 400 | no | 6 | 4 | 90 | 104 | 75.679 | 0,414 | 1,155 | aneuploid | aneuploid | CONCORDANT | 1 | 0 | 0 | 0 | +10, +X | +10, +X | CONCORDANT | | 2 | 21 | 0 | 0 |
| Pb17 | 460 | no | 7 | 4 | 90 | 64 | 53.388 | 0,473 | 1,245 | aneuploid | aneuploid | CONCORDANT | 1 | 0 | 0 | 0 | -14, +20 | -14, +20 | CONCORDANT | | 2 | 21 | 0 | 0 |
| Pb18 | 387 | yes | 3 | EN2 | 100 | 142 | 172.952 | 0,344 | 1,012 | aneuploid | aneuploid | CONCORDANT | 1 | 0 | 0 | 0 | multiple (+1, +4, -5, +17, -21, +22) | multiple (+1, +4, -5, -21, +22) | CONCORDANT | | 5 | 17 | 0 | 1 |
| Pb19 | 400 | no | 9 | 4 | 90 | 60 | 46.297 | 0,514 | 1,404 | aneuploid | aneuploid | CONCORDANT | 1 | 0 | 0 | 0 | +16, -18, -20 | +16, -18, -20 | CONCORDANT | | 3 | 20 | 0 | 0 |
| Pb20 | 453 | yes | 8 | EN3 | 100 | 176 | 233.128 | 0,482 | 1,257 | aneuploid | aneuploid | CONCORDANT | 1 | 0 | 0 | 0 | -7, +16 | multiple (-7, +9, +10, +16, +17) | DISCORDANT | | 2 | 18 | 3 | 0 |
| Pb21 | 281 | yes | 9 | EN3 | 100 | 148 | 197.323 | 0,339 | 0,924 | aneuploid | aneuploid | CONCORDANT | 1 | 0 | 0 | 0 | +19, -21, -22 | -21 | DISCORDANT | | 1 | 20 | 0 | 2 |
| Pb22 | 340 | no | 2 | 5 | 120 | 112 | 97.165 | 0,425 | 1,173 | aneuploid | aneuploid | CONCORDANT | 1 | 0 | 0 | 0 | +12, -13, -16 | +12, -13, -16 | CONCORDANT | | 0 | 23 | 0 | 0 |
| Pb23 | 353 | no | 3 | 5 | 120 | 99 | 82.572 | 0,405 | 1,095 | euploid | euploid | CONCORDANT | 0 | 1 | 0 | 0 | - | - | CONCORDANT | ok | 0 | 23 | 0 | 0 |
| Pb24 | 300 | no | 4 | 5 | 120 | 112 | 92.960 | 0,426 | 1,149 | euploid | euploid | CONCORDANT | 0 | 1 | 0 | 0 | - | - | CONCORDANT | ok | 0 | 23 | 0 | 0 |
| Pb25 | 258 | yes | 1 | EN1 | 120 | 211 | 231.486 | 0,319 | 0,849 | euploid | euploid | CONCORDANT | 0 | 1 | 0 | 0 | - | - | CONCORDANT | ok | 0 | 23 | 0 | 0 |
| Pb26 | 223 | yes | 2 | EN1 | 120 | 281 | 361.988 | 0,277 | 0,750 | aneuploid | aneuploid | CONCORDANT | 1 | 0 | 0 | 0 | -16 | -16 | CONCORDANT | | 1 | 22 | 0 | 0 |
| Pb27 | 295 | yes | 3 | EN1 | 120 | 227 | 268.420 | 0,285 | 0,768 | euploid | euploid | CONCORDANT | 0 | 1 | 0 | 0 | - | - | CONCORDANT | ok | 0 | 23 | 0 | 0 |
| Pb28 | 325 | yes | 4 | EN1 | 120 | 253 | 311.201 | 0,292 | 0,720 | aneuploid | aneuploid | CONCORDANT | 1 | 0 | 0 | 0 | -6, -12 | -6, -12 | CONCORDANT | | 2 | 21 | 0 | 0 |
| Pb29 | 309 | yes | 7 | EN5 | 90 | 183 | 222.899 | 0,307 | 0,813 | euploid | euploid | CONCORDANT | 0 | 1 | 0 | 0 | - | - | CONCORDANT | ok | 0 | 23 | 0 | 0 |
| Pb30 | 331 | yes | 10 | EN3 | 100 | 155 | 220.552 | 0,230 | 0,876 | aneuploid | aneuploid | CONCORDANT | 1 | 0 | 0 | 0 | +11, +15, +22 | +11, +15, +22 | CONCORDANT | | 3 | 20 | 0 | 0 |
| Pb31 | 333 | no | 1 | 7 | 70 | 106 | 75.428 | 0,384 | 1,089 | aneuploid | aneuploid | CONCORDANT | 1 | 0 | 0 | 0 | -17 | -17 | CONCORDANT | | 1 | 22 | 0 | 0 |
| Pb32 | 413 | yes | 4 | EN2 | 100 | 189 | 218.267 | 0,298 | 0,894 | aneuploid | aneuploid | CONCORDANT | 1 | 0 | 0 | 0 | multiple (+3, +11, +12, +16, +19) | multiple (+3, +4, +11, +12, +16, +19) | CONCORDANT | | 5 | 17 | 1 | 0 |
| Pb33 | 413 | yes | 11 | EN3 | 100 | 174 | 243.733 | 0,320 | 0,857 | aneuploid | aneuploid | CONCORDANT | 1 | 0 | 0 | 0 | multiple (-2, +9, -15, +19) | multiple (-2, +9, -15, +19) | CONCORDANT | | 4 | 19 | 0 | 0 |
| Pb34 | 367 | yes | 12 | EN3 | 100 | 132 | 208.681 | 0,301 | 0,824 | aneuploid | aneuploid | CONCORDANT | 1 | 0 | 0 | 0 | -13, -15, -21 | -13, -15, -21 | CONCORDANT | | 3 | 20 | 0 | 0 |
| Pb35 | 331 | no | 5 | 7 | 70 | 128 | 91.228 | 0,362 | 0,978 | aneuploid | aneuploid | CONCORDANT | 1 | 0 | 0 | 0 | -22 | -22 | CONCORDANT | | 1 | 22 | 0 | 0 |
| Pb36 | 360 | no | 6 | 8 | 70 | 108 | 83.115 | 0,366 | 1,020 | euploid | euploid | CONCORDANT | 0 | 1 | 0 | 0 | - | - | CONCORDANT | ok, (+19?) | 0 | 23 | 0 | 0 |
| Pb37 | 333 | no | 7 | 8 | 70 | 94 | 69.548 | 0,489 | 1,317 | aneuploid | aneuploid | CONCORDANT | 1 | 0 | 0 | 0 | +16 | +16 | CONCORDANT | | 1 | 22 | 0 | 0 |
| Pb38 | 300 | yes | 8 | EN5 | 90 | 190 | 246.494 | 0,448 | 1,122 | euploid | euploid | CONCORDANT | 0 | 1 | 0 | 0 | - | - | CONCORDANT | ok, (+22?) | 0 | 23 | 0 | 0 |
| Pb39 | 347 | no | 9 | 8 | 70 | 71 | 54.614 | 0,428 | 1,143 | aneuploid | aneuploid | CONCORDANT | 1 | 0 | 0 | 0 | multiple (-7, +16, -18, -22) | multiple (-7, +16, -18, -22) | CONCORDANT | | 4 | 19 | 0 | 0 |
| Pb40 | 373 | no | 10 | 8 | 70 | 81 | 58.109 | 0,417 | 1,140 | aneuploid | aneuploid | CONCORDANT | 1 | 0 | 0 | 0 | +4 | +4 | CONCORDANT | | 1 | 22 | 0 | 0 |
| Pb41 | 297 | no | 11 | 8 | 70 | 92 | 68.813 | 0,393 | 0,861 | aneuploid | aneuploid | CONCORDANT | 1 | 0 | 0 | 0 | multiple (+3, -6, -7, -8, -9, -10 ,-12, +13, +14, -17, -18, -19, -22) | multiple (+3, -6, -7, -8, +9, -10, -12, +13, +14, -17, -18, -19, -21) | CONCORDANT | | 12 | 9 | 1 | 1 |
| Pb42 | 353 | no | 1 | 9 | 90 | 95 | 65.990 | 0,513 | 1,434 | aneuploid | aneuploid | CONCORDANT | 1 | 0 | 0 | 0 | multiple (+1, +4, +8, +10, +11, +12, +13, +14, -15, +16, +18, +21, +22) | multiple (+1, +4, +8, +10, +11, +12, +14, -15, +16, +18, +22) | CONCORDANT | | 11 | 10 | 0 | 2 |
| Pb43 | 353 | no | 2 | 9 | 90 | 90 | 59.374 | 0,424 | 1,095 | aneuploid | aneuploid | CONCORDANT | 1 | 0 | 0 | 0 | multiple (-4, -7, -8, -10, -12, -13, -17, +19, -20, -21, +22, +X) | multiple (-4, -7, -8, -10, -12, -13, -16, -17, -20, -21) | CONCORDANT | | 9 | 10 | 1 | 3 |
| Pb44 | 433 | no | 3 | 9 | 90 | 73 | 51.420 | 0,455 | 1,302 | aneuploid | aneuploid | CONCORDANT | 1 | 0 | 0 | 0 | +15 | +15 | CONCORDANT | | 1 | 22 | 0 | 0 |
| Pb45 | 347 | no | 4 | 9 | 90 | 102 | 68.937 | 0,401 | 1,128 | aneuploid | aneuploid | CONCORDANT | 1 | 0 | 0 | 0 | +16, -19 | +16, -19 | CONCORDANT | | 2 | 21 | 0 | 0 |
| Pb46 | 347 | no | 5 | 9 | 90 | 100 | 70.807 | 0,408 | 1,125 | aneuploid | euploid | DISCORDANT | 0 | 0 | 0 | 1 | +22 | - | DISCORDANT | ok, (+22?) | 0 | 22 | 0 | 1 |
| Pb47 | 380 | no | 6 | 9 | 90 | 96 | 70.984 | 0,402 | 1,157 | aneuploid | aneuploid | CONCORDANT | 1 | 0 | 0 | 0 | multiple (+10, -14, +16, +18) | multiple (+10, -14, +16, +18) | CONCORDANT | | 4 | 19 | 0 | 0 |
| Pb48 | 323 | no | 7 | 10 | 80 | 35 | 22.063 | 0,614 | 1,809 | euploid | not evaluable | EXCLUDED | | | | | | | EXCLUDED | | | | | |
| Pb49 | 330 | no | 8 | 10 | 80 | 47 | 31.641 | 0,512 | 1,380 | euploid | euploid | CONCORDANT | 0 | 1 | 0 | 0 | - | - | CONCORDANT | ok | 0 | 23 | 0 | 0 |
| Pb50 | 360 | no | 9 | 10 | 80 | 45 | 33.090 | 0,580 | 1,617 | euploid | euploid | CONCORDANT | 0 | 1 | 0 | 0 | - | - | CONCORDANT | ok | 0 | 23 | 0 | 0 |
| Pb51 | 329 | no | 10 | 10 | 80 | 46 | 32.845 | 0,531 | 1,458 | aneuploid | aneuploid | CONCORDANT | 1 | 0 | 0 | 0 | multiple (-7, -9, +16, -19) | multiple (-7, -9, +16, -19) | CONCORDANT | | 4 | 19 | 0 | 0 |
| Pb52 | 321 | no | 11 | 10 | 80 | 52 | 37.722 | 0,480 | 1,236 | euploid | euploid | CONCORDANT | 0 | 1 | 0 | 0 | - | - | CONCORDANT | ok | 0 | 23 | 0 | 0 |
| Pb53 | 380 | no | 12 | 10 | 80 | 58 | 43.439 | 0,576 | 1,587 | euploid | euploid | CONCORDANT | 0 | 1 | 0 | 0 | - | - | CONCORDANT | ok | 0 | 23 | 0 | 0 |
| Pb54 | 302 | no | 1 | 11 | 80 | 54 | 45.582 | 0,477 | 1,308 | aneuploid | aneuploid | CONCORDANT | 1 | 0 | 0 | 0 | -9, -15 | -9, -15 | CONCORDANT | | 2 | 21 | 0 | 0 |
| Pb55 | 413 | no | 2 | 11 | 80 | 51 | 39.452 | 0,484 | 1,329 | aneuploid | aneuploid | CONCORDANT | 1 | 0 | 0 | 0 | -17, -18 | -17, -18 | CONCORDANT | | 2 | 21 | 0 | 0 |
| Pb56 | 367 | no | 3 | 11 | 80 | 51 | 42.753 | 0,526 | 1,425 | aneuploid | aneuploid | CONCORDANT | 1 | 0 | 0 | 0 | -16 | -16 | CONCORDANT | | 1 | 22 | 0 | 0 |
| Pb57 | 380 | no | 4 | 11 | 80 | 62 | 48.039 | 0,463 | 1,269 | euploid | euploid | CONCORDANT | 0 | 1 | 0 | 0 | - | - | CONCORDANT | ok, (+16?) | 0 | 23 | 0 | 0 |
| Pb58 | 353 | yes | 2 | EN4 | 90 | 152 | 96.024 | 0,350 | 0,889 | aneuploid | aneuploid | CONCORDANT | 1 | 0 | 0 | 0 | -13, -14, +17 | multiple (+11, -13, -14, +15, +17, +21) | DISCORDANT | | 3 | 17 | 3 | 0 |
| Pb59 | 360 | no | 6 | 11 | 80 | 40 | 30.948 | 0,526 | 1,539 | aneuploid | aneuploid | CONCORDANT | 1 | 0 | 0 | 0 | multiple (+3, +9, -13, +17, -18, -X) | multiple (+3, +9, -13, +17, -18, -X) | CONCORDANT | | 6 | 17 | 0 | 0 |
| Pb60 | 311 | no | 12 | 12 | 77 | 130 | 35.414 | 0,512 | 1,478 | euploid | euploid | CONCORDANT | 0 | 1 | 0 | 0 | - | - | CONCORDANT | ok | 0 | 23 | 0 | 0 |
| Pb61 | 228 | no | 1 | 12 | 77 | 180 | 50.758 | 0,724 | 1,650 | not evaluable | euploid | EXCLUDED | | | | | | - | EXCLUDED | ok | | | | |
| Pb62 | 340 | yes | 9 | EN5 | 90 | 270 | 171.404 | 0,325 | 0,867 | aneuploid | euploid | DISCORDANT | 0 | 0 | 0 | 1 | +19 | - | DISCORDANT | ok, (+19?) | 0 | 22 | 0 | 1 |
| Pb63 | 373 | no | 3 | 12 | 77 | 180 | 74.607 | 0,508 | 1,317 | aneuploid | aneuploid | CONCORDANT | 1 | 0 | 0 | 0 | multiple (+5, -11, +15, +17, +19) | multiple (+5, -11, +15, +17(s), +19) | CONCORDANT | | 5 | 18 | 0 | 0 |
| Pb64 | 319 | no | 4 | 12 | 77 | 230 | 57.518 | 0,413 | 1,113 | aneuploid | aneuploid | CONCORDANT | 1 | 0 | 0 | 0 | +4 | +4 | CONCORDANT | | 1 | 22 | 0 | 0 |
| Pb65 | 387 | no | 5 | 12 | 77 | 180 | 47.973 | 0,461 | 1,263 | aneuploid | aneuploid | CONCORDANT | 1 | 0 | 0 | 0 | +16 | +16 | CONCORDANT | | 1 | 22 | 0 | 0 |
| Pb66 | 254 | no | 6 | 12 | 77 | 200 | 56.133 | 0,506 | 1,412 | aneuploid | aneuploid | CONCORDANT | 1 | 0 | 0 | 0 | multiple (-9, +10, +11, -13, -16, -18, -20, +21, +22) | multiple (+11, -13, -16, -18, -20, +22) | CONCORDANT | | 6 | 14 | 0 | 3 |
| Pb67 | 371 | no | 1 | 13 | 80 | 56 | 35.184 | 0,500 | 1,372 | aneuploid | aneuploid | CONCORDANT | 1 | 0 | 0 | 0 | +12, -15 | +12, -15 | CONCORDANT | | 2 | 21 | 0 | 0 |
| Pb68 | 301 | no | 2 | 13 | 80 | 77 | 47.042 | 0,431 | 1,200 | aneuploid | aneuploid | CONCORDANT | 1 | 0 | 0 | 0 | +2 | +2 | CONCORDANT | | 1 | 22 | 0 | 0 |
| Pb69 | 356 | no | 3 | 13 | 80 | 50 | 30.943 | 0,560 | 1,538 | aneuploid | aneuploid | CONCORDANT | 1 | 0 | 0 | 0 | +6, -18 | +6, -18 | CONCORDANT | | 2 | 21 | 0 | 0 |
| Pb70 | 294 | no | 4 | 13 | 80 | 71 | 46.296 | 0,431 | 1,182 | aneuploid | aneuploid | CONCORDANT | 1 | 0 | 0 | 0 | -16, -22 | -16, -22 | CONCORDANT | | 2 | 21 | 0 | 0 |
| Pb71 | 353 | no | 5 | 13 | 80 | 63 | 43.959 | 0,465 | 1,290 | aneuploid | aneuploid | CONCORDANT | 1 | 0 | 0 | 0 | +11, +16, -17 | +11, +16, -17 | CONCORDANT | | 3 | 20 | 0 | 0 |
| Pb72 | 445 | no | 6 | 13 | 80 | 75 | 53.594 | 0,484 | 1,218 | aneuploid | aneuploid | CONCORDANT | 1 | 0 | 0 | 0 | -19 | -19 | CONCORDANT | | 1 | 22 | 0 | 0 |
| Pb73 | 510 | no | 7 | 14 | 90 | 53 | 33.809 | 0,484 | 1,390 | aneuploid | aneuploid | CONCORDANT | 1 | 0 | 0 | 0 | +4, -20 | +4, -20 | CONCORDANT | | 2 | 21 | 0 | 0 |
| Pb74 | 474 | yes | 10 | EN5 | 90 | 149 | 214.091 | 0,304 | 0,837 | aneuploid | aneuploid | CONCORDANT | 1 | 0 | 0 | 0 | -16, +21 | -16, +21 | CONCORDANT | | 2 | 21 | 0 | 0 |
| Pb75 | 369 | no | 9 | 14 | 90 | 55 | 36.950 | 0,514 | 1,431 | aneuploid | aneuploid | CONCORDANT | 1 | 0 | 0 | 0 | +16, +22 | +16, +22 | CONCORDANT | | 2 | 21 | 0 | 0 |
| Pb76 | 481 | no | 10 | 14 | 90 | 45 | 28.887 | 0,539 | 1,506 | euploid | euploid | CONCORDANT | 0 | 1 | 0 | 0 | - | - | CONCORDANT | ok | 0 | 23 | 0 | 0 |
| Pb77 | 464 | no | 11 | 14 | 90 | 83 | 55.634 | 0,429 | 1,182 | euploid | euploid | CONCORDANT | 0 | 1 | 0 | 0 | - | - | CONCORDANT | ok | 0 | 23 | 0 | 0 |
| Pb78 | 530 | yes | 3 | EN4 | 90 | 115 | 73.728 | 0,423 | 1,116 | aneuploid | aneuploid | CONCORDANT | 1 | 0 | 0 | 0 | +17, -22 | +17, -22 | CONCORDANT | | 2 | 21 | 0 | 0 |
| Pb79 | 407 | no | 7 | 15 | 100 | 50 | 40.408 | 0,575 | 1,554 | aneuploid | aneuploid | CONCORDANT | 1 | 0 | 0 | 0 | +18 | +18 | CONCORDANT | | 1 | 22 | 0 | 0 |
| Pb80 | 520 | no | 8 | 15 | 100 | 83 | 64.469 | 0,397 | 1,062 | euploid | euploid | CONCORDANT | 0 | 1 | 0 | 0 | - | - | CONCORDANT | ok | 0 | 23 | 0 | 0 |
| Pb81 | 730 | no | 9 | 15 | 100 | 70 | 58.154 | 0,463 | 1,286 | euploid | euploid | CONCORDANT | 0 | 1 | 0 | 0 | - | - | CONCORDANT | ok | 0 | 23 | 0 | 0 |
| Pb82 | 495 | no | 10 | 15 | 100 | 48 | 39.720 | 0,497 | 1,347 | aneuploid | aneuploid | CONCORDANT | 1 | 0 | 0 | 0 | +9, -13 | +9, -13 | CONCORDANT | | 2 | 21 | 0 | 0 |
| Pb83 | 550 | no | 11 | 15 | 100 | 76 | 62.625 | 0,418 | 1,122 | aneuploid | aneuploid | CONCORDANT | 1 | 0 | 0 | 0 | +16, -19 | +16, -19 | CONCORDANT | | 2 | 21 | 0 | 0 |
| Pb84 | 479 | no | 12 | 15 | 100 | 54 | 44.664 | 0,519 | 1,345 | aneuploid | aneuploid | CONCORDANT | 1 | 0 | 0 | 0 | +1 | +1 | CONCORDANT | | 1 | 22 | 0 | 0 |
| Pb85 | 520 | yes | 11 | EN5 | 90 | 155 | 246.406 | 0,330 | 0,870 | aneuploid | euploid | DISCORDANT | 0 | 0 | 0 | 1 | -16 | - | DISCORDANT | ok, (-16?) | 0 | 22 | 0 | 1 |
| Pb86 | 411 | yes | 5 | EN2 | 100 | 261 | 330.514 | 0,427 | 1,062 | aneuploid | aneuploid | CONCORDANT | 1 | 0 | 0 | 0 | multiple (-6q, -8, -9q, -part.15, +16p, -17, +19, +22) | multiple (-8, -9, -14, -17, +22) | CONCORDANT | | 3 | 14 | 1 | 1 |
| Pb87 | 680 | yes | 6 | EN2 | 100 | 158 | 185.739 | 0,334 | 0,900 | aneuploid | aneuploid | CONCORDANT | 1 | 0 | 0 | 0 | +20 | +20 | CONCORDANT | | 1 | 22 | 0 | 0 |
| Pb88 | 640 | no | 10 | 16 | 100 | 37 | 25.588 | 0,588 | 1,650 | aneuploid | aneuploid | CONCORDANT | 1 | 0 | 0 | 0 | -17 | -17 | CONCORDANT | | 1 | 22 | 0 | 0 |
| Pb89 | 640 | no | 11 | 16 | 100 | 53 | 36.176 | 0,503 | 1,357 | aneuploid | aneuploid | CONCORDANT | 1 | 0 | 0 | 0 | +22 | +22 | CONCORDANT | | 1 | 22 | 0 | 0 |
| Pb90 | 700 | no | 12 | 16 | 100 | 40 | 27.638 | 0,562 | 1,512 | aneuploid | aneuploid | CONCORDANT | 1 | 0 | 0 | 0 | multiple (-4, -7, +16, -19) | multiple (-4, -7, +16, -19) | CONCORDANT | | 4 | 19 | 0 | 0 |
| Pb91 | 690 | yes | 4 | EN4 | 90 | 28 | 92.844 | 0,357 | 0,915 | aneuploid | aneuploid | CONCORDANT | 1 | 0 | 0 | 0 | -2, +6 | -2, +6 | CONCORDANT | | 2 | 21 | 0 | 0 |
| Pb92 | 630 | no | 2 | 17 | 80 | 61 | 48.297 | 0,472 | 1,197 | aneuploid | aneuploid | CONCORDANT | 1 | 0 | 0 | 0 | multiple (-2, -13, -14, +16) | multiple (-2, -13, -14, +16) | CONCORDANT | | 4 | 19 | 0 | 0 |
| Pb93 | 294 | yes | 5 | EN4 | 90 | 47 | 181.363 | 0,315 | 0,792 | aneuploid | aneuploid | CONCORDANT | 1 | 0 | 0 | 0 | +1, -16, +19 | +1, -16 | DISCORDANT | | 2 | 20 | 0 | 1 |
| Pb94 | 288 | no | 4 | 17 | 80 | 59 | 49.543 | 0,436 | 1,179 | euploid | euploid | CONCORDANT | 0 | 1 | 0 | 0 | - | - | CONCORDANT | ok (+18?, -22?) | 0 | 23 | 0 | 0 |
| Pb95 | 338 | yes | 12 | EN5 | 90 | 124 | 187.577 | 0,316 | 0,811 | not evaluable | euploid | EXCLUDED | | | | | | - | EXCLUDED | ok | | | | |
| Pb96 | 252 | yes | 6 | EN4 | 90 | 158 | 138.336 | 0,296 | 0,774 | aneuploid | aneuploid | CONCORDANT | 1 | 0 | 0 | 0 | +11,-16,+19 | +11, -16 | DISCORDANT | | 2 | 20 | 0 | 1 |
| Pb97 | 670 | no | 7 | 18 | 90 | 48 | 36.475 | 0,525 | 1,557 | aneuploid | aneuploid | CONCORDANT | 1 | 0 | 0 | 0 | +16 | +16 | CONCORDANT | | 1 | 22 | 0 | 0 |
| Pb98 | 580 | no | 8 | 18 | 90 | 73 | 54.231 | 0,448 | 1,155 | aneuploid | aneuploid | CONCORDANT | 1 | 0 | 0 | 0 | -4 | -4 | CONCORDANT | | 1 | 22 | 0 | 0 |
| Pb99 | 580 | no | 9 | 18 | 90 | 53 | 40.400 | 0,564 | 1,495 | aneuploid | aneuploid | CONCORDANT | 1 | 0 | 0 | 0 | +16 | +16 | CONCORDANT | | 1 | 22 | 0 | 0 |
| Pb100 | 438 | no | 10 | 18 | 90 | 56 | 45.815 | 0,503 | 1,350 | euploid | euploid | CONCORDANT | 0 | 1 | 0 | 0 | - | - | CONCORDANT | ok | 0 | 23 | 0 | 0 |
| Pb101 | 444 | no | 11 | 18 | 90 | 67 | 50.564 | 0,475 | 1,293 | euploid | euploid | CONCORDANT | 0 | 1 | 0 | 0 | - | - | CONCORDANT | ok, (+22?) | 0 | 23 | 0 | 0 |
| Pb102 | 444 | no | 12 | 18 | 90 | 49 | 38.188 | 0,489 | 1,354 | euploid | euploid | CONCORDANT | 0 | 1 | 0 | 0 | - | - | CONCORDANT | ok | 0 | 23 | 0 | 0 |
