## Supplementary Table S2 for "Evaluation of nanopore sequencing on polar bodies for routine pre-implantation genetic testing for aneuploidy"

| Sample | Run | | Time / Sample (min) | | Yield [Mb] | | Raw Reads | | noise | | MAPD | | Ploidy Status | | | | | Chromosome aberrations | | | | |
|---|---|---|---|---|---|---|---|---|---|---|---|---|---|---|---|---|---|---|---|---|---|---|
| Sample Number | Without EN | With EN | Without EN | With EN | Without EN | With EN | Without EN | With EN | Without EN | With EN | Without EN | With EN | aCGH | Nanopore | Nanopore with EN | Concordance without EN | Concordance with EN | aCGH | Nanopore | Nanopore with EN | Concordance without EN | Concordance with EN |
| Pb25 | 6 | EN1 | 96 | 120 | 102 | 211 | 77.207 | 231.486 | 0,369 | 0,319 | 0,963 | 0,849 | euploid | euploid | euploid | CONCORDANT | CONCORDANT | - | - | - | CONCORDANT | CONCORDANT |
| Pb26 | 6 | EN1 | 96 | 120 | 118 | 281 | 87.497 | 361.988 | 0,378 | 0,277 | 1,033 | 0,75 | aneuploid | aneuploid | aneuploid | CONCORDANT | CONCORDANT | -16 | -16 | -16 | CONCORDANT | CONCORDANT |
| Pb27 | 6 | EN1 | 96 | 120 | 101 | 227 | 71.566 | 268.420 | 0,402 | 0,285 | 1,152 | 0,768 | euploid | euploid | euploid | CONCORDANT | CONCORDANT | - | - | - | CONCORDANT | CONCORDANT |
| Pb28 | 6 | EN1 | 96 | 120 | 109 | 253 | 86.791 | 311.201 | 0,395 | 0,292 | 1,047 | 0,72 | aneuploid | aneuploid | aneuploid | CONCORDANT | CONCORDANT | -6, -12 | -6, -12 | -6, -12 | CONCORDANT | CONCORDANT |
| Pb10 | 3 | EN2 | 70 | 100 | 67 | 178 | 59.844 | 189.572 | 0,431 | 0,324 | 1,201 | 0,894 | euploid | euploid | euploid | CONCORDANT | CONCORDANT | - | - | - | CONCORDANT | CONCORDANT |
| Pb12 | 3 | EN2 | 90 | 100 | 58 | 245 | 46.745 | 274.533 | 0,394 | 0,281 | 1,149 | 0,771 | euploid | euploid | euploid | CONCORDANT | CONCORDANT | - | - | - | CONCORDANT | CONCORDANT |
| Pb18 | 4 | EN2 | 90 | 100 | 195 | 142 | 164.091 | 172.952 | 0,351 | 0,344 | 0,950 | 1,012 | aneuploid | aneuploid | aneuploid | CONCORDANT | CONCORDANT | multiple | multiple | multiple | CONCORDANT | CONCORDANT |
| Pb32 | 7 | EN2 | 70 | 100 | 109 | 189 | 83.677 | 218.267 | 0,397 | 0,298 | 1,134 | 0,894 | aneuploid | aneuploid | aneuploid | CONCORDANT | CONCORDANT | multiple | multiple | multiple | CONCORDANT | CONCORDANT |
| Pb86 | 16 | EN2 | 100 | 100 | 101 | 261 | 67.671 | 330.514 | 0,500 | 0,427 | 1,244 | 1,062 | aneuploid | aneuploid | aneuploid | CONCORDANT | CONCORDANT | multiple | multiple | multiple | CONCORDANT | CONCORDANT |
| Pb87 | 16 | EN2 | 100 | 100 | 49 | 158 | 34.733 | 185.739 | 0,516 | 0,334 | 1,479 | 0,900 | aneuploid | aneuploid | aneuploid | CONCORDANT | CONCORDANT | +20 | +20 | +20 | CONCORDANT | CONCORDANT |
| Pb14 | 3 | EN3 | 90 | 100 | 68 | 144 | 54.967 | 190.505 | 0,449 | 0,328 | 1,272 | 0,896 | euploid | euploid | euploid | CONCORDANT | CONCORDANT | - | - | - | CONCORDANT | CONCORDANT |
| Pb20 | 4 | EN3 | 90 | 100 | 105 | 176 | 92.290 | 233.128 | 0,537 | 0,482 | 1,389 | 1,257 | aneuploid | aneuploid | aneuploid | CONCORDANT | CONCORDANT | -7, +16 | multiple | multiple | DISCORDANT | DISCORDANT |
| Pb21 | 5 | EN3 | 120 | 100 | 92 | 148 | 74.701 | 197.323 | 0,388 | 0,339 | 1,074 | 0,924 | aneuploid | aneuploid | aneuploid | CONCORDANT | CONCORDANT | +19, -21, -22 | -21 | -21 | DISCORDANT | DISCORDANT |
| Pb30 | 7 | EN3 | 70 | 100 | 96 | 155 | 69.559 | 220.552 | 0,390 | 0,320 | 1,104 | 0,876 | aneuploid | aneuploid | aneuploid | CONCORDANT | CONCORDANT | +11, +15, +22 | +11, +15 | +11, +15, +22 | DISCORDANT | CONCORDANT |
| Pb33 | 7 | EN3 | 70 | 100 | 86 | 174 | 62.702 | 243.733 | 0,427 | 0,320 | 1,140 | 0,857 | aneuploid | aneuploid | aneuploid | CONCORDANT | CONCORDANT | multiple | -2, +9, -15 | +2, +9, -15 | DISCORDANT | DISCORDANT |
| Pb34 | 7 | EN3 | 70 | 100 | 106 | 132 | 78.244 | 208.681 | 0,378 | 0,301 | 1,047 | 0,824 | aneuploid | aneuploid | aneuploid | CONCORDANT | CONCORDANT | -13,-15,-21 | -13, -15 | -13,-15,-21 | DISCORDANT | CONCORDANT |
| Pb6 | 2 | EN4 | 70 | 90 | 86 | 170 | 79.915 | 117.144 | 0,337 | 0,339 | 1,005 | 0,927 | euploid | euploid | euploid | CONCORDANT | CONCORDANT | - | - | - | CONCORDANT | CONCORDANT |
| Pb58 | 11 | EN4 | 80 | 90 | 79 | 152 | 65.709 | 96.024 | 0,398 | 0,350 | 1,041 | 0,889 | aneuploid | aneuploid | aneuploid | CONCORDANT | CONCORDANT | -13,-14,+17 | multiple | multiple | DISCORDANT | DISCORDANT |
| Pb78 | 14 | EN4 | 90 | 90 | 72 | 115 | 48.876 | 73.728 | 0,503 | 0,423 | 1,368 | 1,116 | aneuploid | aneuploid | aneuploid | CONCORDANT | CONCORDANT | +17,-22 | +17 | +17,-22 | DISCORDANT | CONCORDANT |
| Pb91 | 17 | EN4 | 80 | 90 | 28 | 28 | 21.405 | 92.844 | 0,630 | 0,357 | 1,671 | 0,915 | aneuploid | aneuploid | aneuploid | CONCORDANT | CONCORDANT | -2,+6 | -2, -21 | -2,+6 | DISCORDANT | CONCORDANT |
| Pb93 | 17 | EN4 | 80 | 90 | 47 | 47 | 44.624 | 181.363 | 0,442 | 0,315 | 1,173 | 0,792 | aneuploid | aneuploid | aneuploid | CONCORDANT | CONCORDANT | +1,-16,+19 | +1, -16 | +1, -16 | DISCORDANT | DISCORDANT |
| Pb96 | 17 | EN4 | 80 | 90 | 66 | 158 | 60.677 | 138.336 | 0,389 | 0,296 | 1,017 | 0,774 | aneuploid | aneuploid | aneuploid | CONCORDANT | CONCORDANT | +11,-16,+19 | +11, -16 | +11, -16 | DISCORDANT | DISCORDANT |
| Pb29 | 6 | EN5 | 96 | 90 | 98 | 183 | 77.560 | 222.899 | 0,403 | 0,307 | 1,082 | 0,813 | euploid | euploid | euploid | CONCORDANT | CONCORDANT | - | - | - | CONCORDANT | CONCORDANT |
| Pb38 | 8 | EN5 | 70 | 90 | 110 | 190 | 82.209 | 246.494 | 0,506 | 0,448 | 1,266 | 1,122 | euploid | aneuploid | euploid | DISCORDANT | CONCORDANT | - | -17 | - | DISCORDANT | CONCORDANT |
| Pb62 | 12 | EN5 | 77 | 90 | 180 | 136 | 51.856 | 171.404 | 0,441 | 0,325 | 1,155 | 0,867 | aneuploid | euploid | euploid | DISCORDANT | DISCORDANT | +19 | - | - | DISCORDANT | DISCORDANT |
| Pb74 | 14 | EN5 | 90 | 90 | 76 | 149 | 48.009 | 214.091 | 0,447 | 0,304 | 1,233 | 0,837 | aneuploid | euploid | aneuploid | DISCORDANT | CONCORDANT | -16,+21 | - | -16,+21 | DISCORDANT | CONCORDANT |
| Pb85 | 16 | EN5 | 100 | 90 | 61 | 155 | 42.281 | 246.406 | 0,482 | 0,330 | 1,305 | 0,870 | aneuploid | euploid | euploid | DISCORDANT | DISCORDANT | -16 | - | - | DISCORDANT | DISCORDANT |
| Pb95 | 17 | EN5 | 80 | 90 | 50 | 124 | 44.210 | 187.577 | 0,440 | 0,316 | 1,179 | 0,811 | not evaluable | euploid | euploid | EXCLUDED | EXCLUDED | not evaluable | - | - | EXCLUDED | EXCLUDED |
