## Supplementary Table S3 for "Evaluation of nanopore sequencing on polar bodies for routine pre-implantation genetic testing for aneuploidy"

| Step | Material | Cost/sample for 1 sample | Cost/sample for 12 samples | Time for 1 sample | Time for 12 samples |
|---|---|---|---|---|---|
| **WGA** | PicoPlex WGA Kit | 48 € | 26 € | 2,5 h | 2,5 h |
| OR: | | OR | OR | OR | OR |
| in our study: | REPLIg Single Cell Kit incl. Endonuclease I digest | 54 € | 30 € | 2.5 h | 3.5 h |
| OR: | | OR | OR | OR | OR |
| | REPLIg Ultra Fast Mini Kit Incl. Endonuclease I digest | 26 € | 14 € | 1,5 h | 2,5 h |
| | CAVE: One negative control is included in each WGA run to detect contamination and will not be handled further. | | | | |
| **Library Prep** | details see Suppl. Table 3 | 70€ | 45€ | 1 h | 2 x 5 h |
| **Sequencing** (and Basecalling) | Flow Cell (**min. 48 Flow cells/year**) | 95€ → calculated if flow cell is re-used 5x for one sample each | 40€ | 30 min | 2 x 6 h |
| **Data analysis** | PC/NAS/Server | | | 5 min | 30-60 min |
| **TOTAL (dependent on WGA Kit)** | | **190€ - 220€** | **100€ - 120€** | **3 h – 5 h** | **14 h – 16 h** |
