## Supplementary Table S4 for "Evaluation of nanopore sequencing on polar bodies for routine pre-implantation genetic testing for aneuploidy"

**WGA:**

| | | |
|---|---|---|
| ○ PicoPlex WGA Kit (Takara #R30050, 50 rx) | 1188€ | 23,76€/sample |
| OR: | | |
| ○ REPLI-g Ultra Fast Mini Kit (QIAGEN #150035, 100 rx) | 901€ | 9€/sample |
| ○ OR: REPLI-g Single Cell Kit (QIAGEN #150345, 96rx) | 2202€ | 23€/sample |
| ○ NEB T7 Endonuclease I (NEB, #M0302L) | 293€ | 4,20€/sample |

**Library Prep:**

| | | |
|---|---|---|
| ○ NEBNext Ultra II End Prep/dA Tailing Module (NEB #E7546L, 96rx) | 835€ | 8,70€/sample |
| ○ NEBNext FFPE DNA Repair Mix (NEB, #M6630L, 96rx) | 604€ | 6,30€/sample |
| ○ gDNA Clean & Concentrator-25 (Zymo #D4065, 100rx) | 329€ | 3,29€/ sample |
| ○ 2x DNA Clean & Concentrator-5 (Zymo #D4004, 200rx) | 299€ | 2,99€/ sample |
| ○ Elution Buffer (QIAGEN, #19086, 250 ml) | 35€ | 0,50€/ sample |
| ○ Blunt/TA Ligase Master Mix (NEB #M0367, 50rx) | 370€ | 7,40€/ sample |
| ○ Quick T4 DNA Ligase (NEB #M2200L, 15 Libraries) | 396€ | 26€/library |
| ○ Native barcoding kit (ONT, EXP-NBD104, 12 BC) | 259€ | 3,60€/ sample |
| ○ Ligation Sequencing Expansions (instead of SQK-LSK109) (ONT, SFB001, AUX001, AMII001, each 12rx) | 359€ | ca. 4€/ sample |
| ○ Flow Cell Priming Kit (ONT, EXP-FLP002) | 35€ | ca. 1€/ sample |
| ○ Flow Cell Wash Kit (ONT, WSH004) | 90€ | ca. 2€/ sample |

**Quality control:**

| | | |
|---|---|---|
| ○ Agarose E-Gel 2% (Thermo, #A42135, 10 Gels) | 120€ | 1,50€/ sample |
| ○ 3x Qubit BR Assay (Thermo, #Q32853, 500rx) | 327€ | 2€/ sample |

**For one sample: 70€ / sample**

**For 12 samples: 45 € / sample**
